# Morbidity and mortality burden of COVID-19 in rural Madagascar: results from a longitudinal cohort and nested seroprevalence study

**DOI:** 10.1101/2023.03.24.23287674

**Authors:** Andres Garchitorena, Lova Tsikiniaina Rasoloharimanana, Rado JL Rakotonanahary, Michelle V Evans, Ann C. Miller, Karen E. Finnegan, Laura F. Cordier, Giovanna Cowley, Benedicte Razafinjato, Marius Randriamanambintsoa, Samuel Andrianambinina, Stephen Popper, Raphaël Hotahiene, Matthew H. Bonds, Matthieu Schoenhals

**Author notes:** Equal contribution (Matthew H. Bonds, Matthieu Schoenhals).

## Abstract

**Introduction:** Three years into the pandemic, there remains significant uncertainty about the true infection and mortality burden of COVID-19 in the WHO-Africa region. High quality, population-representative studies in Africa are rare and tend to be conducted in national capitals or large cities, leaving a substantial gap in our understanding of the impact of COVID-19 in rural, low-resource settings. Here, we estimated the spatio-temporal morbidity and mortality burden associated with COVID-19 in a rural health district of Madagascar until the first half of 2021.

**Methods:** We integrated a nested seroprevalence study within a pre-existing longitudinal cohort conducted in a representative sample of 1600 households in Ifanadiana District, Madagascar. Socio-demographic and health information was collected in combination with dried blood spots for about 6500 individuals of all ages, which were analysed to detect IgG and IgM antibodies against four specific proteins of SARS-CoV2 in bead-based multiplex immunoassay. We evaluated spatio-temporal patterns in COVID-19 infection history and its associations with several geographic, socio-economic and demographic factors via logistic regressions.

**Results:** Eighteen percent of people had been infected by April-June 2021, with seroprevalence increasing with individuals’ age. COVID-19 primarily spread along the only paved road and in major towns during the first epidemic wave, subsequently spreading along secondary roads during the second wave to more remote areas. Wealthier individuals and those with occupations such as commerce and formal employment were at higher risk of being infected in the first wave. Adult mortality increased in 2020, particularly for older men for whom it nearly doubled up to nearly 40 deaths per 1000. Less than 10% of mortality in this period could be directly attributed to COVID-19 deaths given known infection fatality ratios and observed seroprevalence in the district.

**Conclusion:** Our study provides a very granular understanding on COVID-19 transmission and mortality in a rural population of sub-Saharan Africa and suggests that the disease burden in these areas may have been substantially underestimated.

## Introduction

Soon after the initial outbreak of COVID-19 in Wuhan, China, scientists estimated the key epidemiological properties that determined the spread and impact of the disease, predicting a global pandemic that would ultimately infect the majority of people in the world and kill millions [1–3]. By the end of 2021, an estimated 3.39 billion people globally had been infected and over 18 million had died [4,5]. As testing capacity increased and the pandemic progressed, the role of age, gender, and comorbidities as risk factors became clearer [6], but key aspects of the epidemic remained uncertain for areas of the world where high-quality data was not available, leading to considerable debate about the expected burden in these areas [7,8]. This was especially pronounced and persistent in the World Health Organization (WHO) Africa region. At the onset of the pandemic, there was substantial concern that the region could be especially vulnerable for the same reasons it suffers from high burdens of other infectious diseases, low access to health care and limited capacity for treatment [9,10]. As the pandemic continued, low rates of reported cases and deaths corresponded to a growing chorus suggesting that perhaps Africa was at lower risk for COVID-19 morbidity and mortality due to genetic, environmental, or immunological differences [11,12].

Three years on, there remains significant uncertainty about the true infection and mortality burden of COVID-19 in the WHO-Africa region. Low capacity for routine clinical testing of COVID-19 cases has undermined the use of government statistics for understanding COVID-19 spread. Several recent seroprevalence studies have demonstrated that patterns of infection in Africa are consistent with patterns observed elsewhere, ranging between 40 and 60% positivity by mid-2021 [13]. In fact, sub-Saharan Africa could be the region with the highest infection rates globally [4]. However, high quality, representative studies in Africa continue to be rare. Studies tend to rely on opportunistic data collection, such as from blood donors [14–19], healthcare workers [20–25], or patients coming to health facilities for reasons other than COVID-19 [26–31], none of which are demographically representative of the general population. In addition, where population-representative seroprevalence studies do exist, they tend to be conducted in national capitals or large cities, and the majority have a moderate to high risk of bias (e.g. tests used, sampling design, etc.) [13].

A second major challenge in the WHO Africa region is estimating the mortality burden associated with COVID-19 from existing sources. Indeed, the limited availability of vital registration records means that deaths are not systematically reported [32], and higher rates of background mortality in these populations can obscure the characterization of excess deaths from COVID-19 [33]. In addition, the indirect death toll of an epidemic can be substantial due to health system disruptions, lower access to healthcare and other factors, as observed during the 2014-2015 Ebola epidemic [34,35]. Consequently, estimates of the COVID-19 mortality burden for the African Region vary widely. For instance, while the WHO estimates that less than 500,000 died from COVID-19 in 2020-2021 using data from reported case fatality rates and seroprevalence studies [36], other studies suggest this number could be three to four times higher [4,5,37]. All these challenges are exacerbated in poor, rural areas of sub-Saharan Africa, where there is virtually no quality, population-representative information on COVID-19 seroprevalence or mortality.

Here, we take advantage of a pre-existing longitudinal cohort study in a representative sample of a health district in rural Madagascar to estimate the morbidity and mortality burden associated with COVID-19 in this population until April-June of 2021. In addition to standard socio-demographic and health information, we collected dried blood spots in about 6500 individuals of all ages, which were analysed to detect IgG and IgM antibodies against four specific proteins of SARS-CoV2 via bead-based multiplex immunoassay. We evaluated patterns in SARS-CoV-2 infection history across space and time in the context of several socio-economic and demographic factors. This resulted in the most granular data on COVID-19 for a rural population of the WHO Africa Region that we are aware of, providing insights into COVID-19 epidemiology for remote rural populations with high rates of poverty.

## Methods

### Study site

Ifanadiana is a rural health district of approximately 200,000 people located in the region of Vatovavy, in Southeastern Madagascar. The vast majority of the district’s population (85%) relies on subsistence agriculture and about three quarters live in extreme poverty [38]. Terrain in the majority of the district consists of deep valleys and a mountainous landscape, while road infrastructure is scarce. A single paved road runs across the district from East to West through four of its communes, and two main non-paved roads connect the district capital, Ifanadiana, with the communes in the North and South. Only a fraction of these roads is accessible by all-terrain vehicles and the availability of such vehicles is limited. The rest of the road network consists of smaller paths accessible by all-terrain motorcycles, with most of the network accessible only by foot.

The district has one reference hospital, one main primary care health center (CSB II) for each of its 15 communes (subdivision of a district with ∼15,000 people), six additional smaller primary health centers (CSB I) for its larger communes, and one community health site with two community health workers for each of its 195 Fokontany (subdivision of a commune with ∼1,000 population). Beginning in 2014, an integrated health-system strengthening (HSS) intervention has been implemented via a partnership between the Madagascar Ministry of Public Health (MoH) and the non-governmental organization Pivot [39]. This intervention is guided by existing MoH policies, covers all six WHO building blocks of HSS, and is implemented across all three levels of care in the district (community, primary health center, and hospital). HSS is done through integration of horizontal improvements in system “readiness”, vertically aligned clinical programs, and information systems at all levels of care. Despite HSS efforts in place, diagnostic rates for COVID-19 were low – similar to much of rural Africa. At the time of the survey less than 350 cases had been reported in the whole Vatovavy region since the beginning of the pandemic [40]. The current study was carried out in support of these HSS efforts, to better understand the burden of COVID-19 in the district during the first two epidemic waves.

### Survey data collection

A seroprevalence survey was added to an existing longitudinal cohort study initiated in 2014 to obtain demographic, health and socio-economic information from a representative sample of 1,600 households in Ifanadiana District over time. Questionnaires in the cohort were mostly adapted from the Demographic and Health Survey [41]. The Madagascar National Institute of Statistics (INSTAT), which implements all major national health surveys in the country, was responsible for data collection, survey coordination, training and oversight. A two-stage sample stratified the district by the HSS intervention’s initial catchment area. Eighty clusters, half from each stratum, were selected at random from enumeration areas mapped during the 2009 census, and households were then mapped within each cluster. Twenty households were selected at random from each cluster.

Four waves of data collection have been conducted in 2014, 2016, 2018 and 2021, in which the original 1600 households were revisited. Any missing households (or those that declined to continue) were replaced with others from the same cluster using a predefined random replacement list. Response rates were about 95% for each wave [42]. Individual face-to-face interviews were conducted with all women aged 15 to 49 years and men aged 15 to 59 years (usual residents or visitors). Data collected in the questionnaires included, among others, household composition (size, genders, ages); indicators of socio-economic status (education, employment, household durable assets); illness in last 30 days; preventive behavior (bed net ownership, access to water and sanitation); women’s reproductive history and care seeking behavior for reproductive health; children’s health, development and care-seeking for illness; adult, maternal and child mortality. To learn more about the impact of the COVID-19 pandemic in this area, the 2021 wave of data collection (April 22^nd^ to June 20^th^) included, for each household member, questions on COVID-related symptoms in the previous 6 months. In addition, for consenting individuals of all ages, a dried blood spot (DBS) was obtained by finger prick using a single-use lancet needle by trained nurses, with 1–5 DBS collected on Whatman 903 Protein Caver Card filter papers for each individual.

French and Malagasy questionnaires used in the cohort, as well as data collection protocols, were standardized and validated for Madagascar during previous national surveys carried out by INSTAT. The study was approved by the Madagascar National Ethics Committee and Harvard Medical School IRB, including amendments for changes in the 2021 surveys. All adults (≥ 15 years) provided oral informed consent for the in-person interview and written informed consent for biological sample collection. Parents or guardians provided written consent for biological sample collection from children ≤ 15 years of age, and children 7-14 years provided written assent separately. To ensure the safety of both the study teams and participants and to avoid transmission of COVID-19 during the survey, field protocols were adapted based on guidance from the SMART initiative for resuming population and household surveys [43]. This included, among others, frequent testing of survey teams before, during and after the survey; initial quarantine of all teams on site before beginning the survey; immediate quarantine of any COVID-positive staff from the survey; use of masks and other protective equipment for survey teams and participants during interviews.

### Serological analyses of dried blood spots

Dried blood spots were cut to obtain 3mm-subpunches that were then shaken overnight at 4°C in PBS-T 0.5% (VWR, E404 and Sigma, P1379). Elutions were then transferred to 1.5ml tubes and stored at - 20°C. A pool of samples of patients recruited before the pandemic (early 2019) was used as a negative control. The positive control was a pool of a panel obtained from the WHO. Using methods previously described [44], but with eluted samples instead of serum, samples were processed using a MBA (multiplex bead assay) on the Luminex platform (MagpixTM) for antibodies against 4 antigens of SARS-CoV2: Spike S1 (GenScript, Z03485), Spike S2 (Sinobiological 40590-V08B), Spike RBD (GenScript, Z03483) and NP (GenScript, Z03480). Magnetic beads (Luminex, MagplexTM MC100XX-01) were coupled to these four antigens using xMAP Antibody coupling kit (Luminex, 40-50016). Briefly, 10 μL of Sulfo-NHS and 10μl EDC were added to 5.106 washed beads and then 25μg of recombinant protein were added to the mixture and incubated for 2 hours in the dark. A multiplex of 1000 beads per antigen was transferred in each well of a 96-well round bottom microplates (Fisher Scientific, Corning 3789). 100μl of non-diluted eluates and controls were added and incubated under agitation during 45 minutes. After two washing steps, 100μl of secondary antibodies IgG (Southern Biotech, Goat Anti-human IgG (FC), 2042-09) or IgM (Thermofisher Scientific, Goat Anti-human IgM (FC) R-PE, H15104) were added to the beads, with shaking during 45 minutes. Finally, the beads were resuspended in 120μl of buffer and analysed on MagpixTM. The signal of the median fluorescence intensity (MFI) was detected by standard PMT.

Cut-off limits for determining positive antibodies for SARS-CoV-2 were based on receiver operator characteristics (ROC) analysis using Graphpad Prism 8.0.1. The samples used in this analysis were 42 Covid-19 negative DBS samples (patients recruited before the pandemic) and 74 COVID-19 positive DBS samples from a cohort of COVID-19 cases followed-up since the beginning of the epidemic in Madagascar (FFX cohort) [45]. Normalization of the results using the positive and the negative control in each plate was carried out to avoid possible batch-effect. A second correction for differing volumes of blood spots differing from one sample to another was performed by correcting the Ig concentration with the total protein concentration measured using a Bradford protein assay and following the manufacturer’s recommendations (Biorad, 500-0006).

### Data analyses

#### Seroprevalence of recent and past SARS-CoV2 infections

To obtain seroprevalence estimates, we carried out two sets of analyses. First, normalised and protein-corrected values of mean fluorescence intensity (MFI) for each of the eight SARS-CoV2 antibodies were compared with their corresponding positivity threshold obtained from the FFX cohort [45] to determine whether the sample was positive or negative for that particular antibody. This resulted in eight different seroprevalence values, one for each antibody considered. Second, to reduce the number of dimensions of the serological data and obtain discrete consistent groups we used k-means clustering to classify individuals’ sero-positivity based on normalized and protein-corrected MFI values from the different antibodies. Data were visually inspected for outliers, and 134 individuals who had abnormally high values of antibodies (outside the range of mean + 3 sd) were removed. We preconditioned the data via a principal components analysis to reduce its dimensionality [46], and used the first two principal components, which explained over 60% of the variance, in the subsequent cluster analysis. We then estimated the optimal number of clusters via the average silhouette width, which identified three as the optimal number of clusters. We used the Hartigan-Wong algorithm to perform k-means clustering using 999 starting sets of centroids.

Clusters were assigned meaningful sero-positivity labels based on the component loadings of the initial PCA and the clusters’ locations relative to these loadings (Figure 1). IgG and IgM antibodies had a strong positive effect on the first principal component (PC1), representing the gradient between overall positive and negative sero-positivity. The second principal component had strong positive loadings for IgM antibodies, which are indicative of recent infection[47], and therefore represented the difference between past and recent infections.

**Figure 1.**
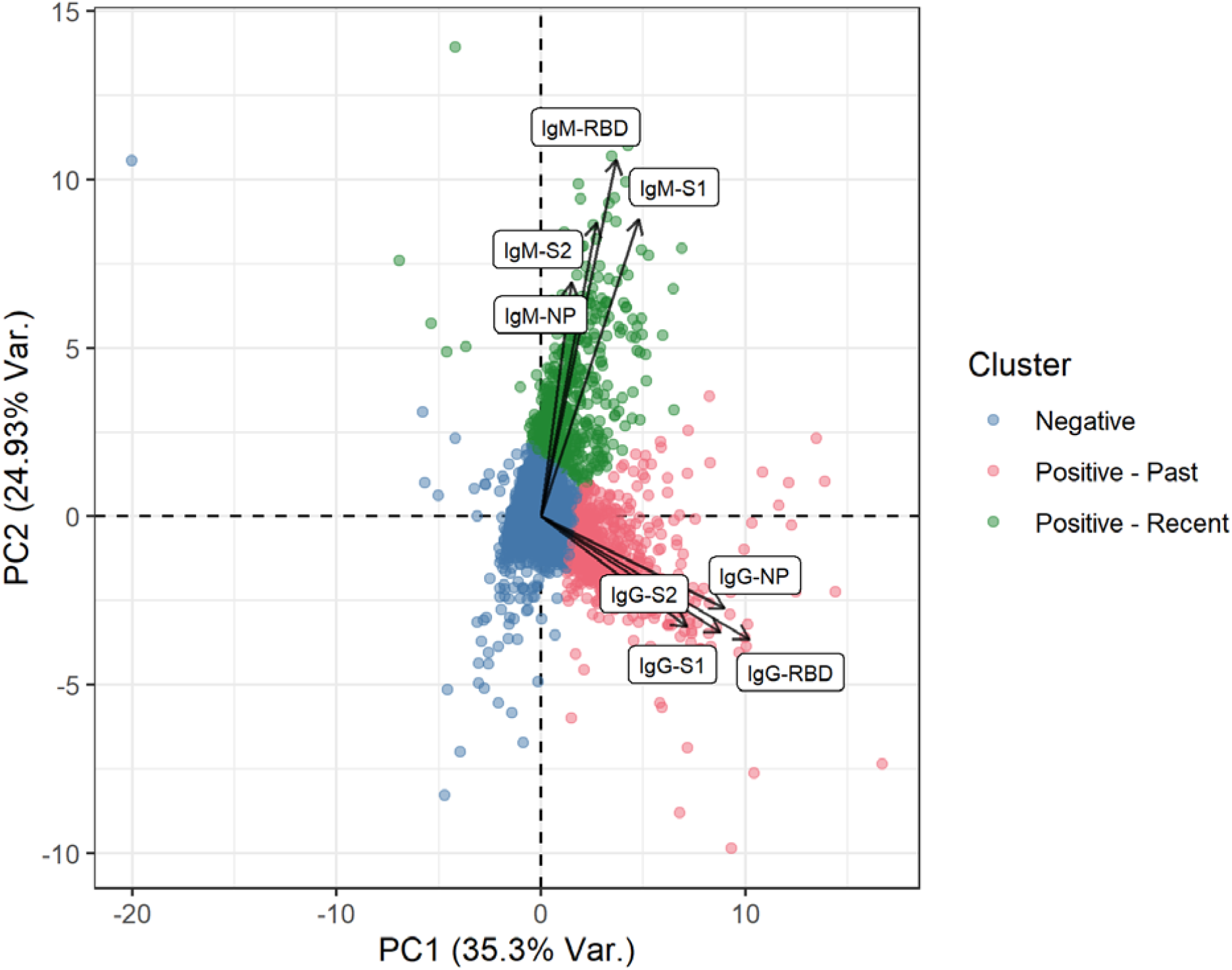
K-means clustering of SARS-CoV2 seroprevalence. Results are based on normalized mean fluorescence intensity values for eight antibodies against SARS-CoV2 (names in white boxes). Colours represent the three clusters obtained, plotted along the axes of the first two components of a principal components analysis that explained over 60% of the variance in these antibodies.

#### Trends and factors associated with SARS-CoV2 seroprevalence

Statistical analyses were carried out to understand local patterns of COVID-19 seroprevalence in Ifanadiana District, including geographic trends as well as associations with socio-economic and demographic factors. First, to study geographic trends in recent and past SARS-CoV2 infections in Ifanadiana, average seroprevalence values for the 80 geographic clusters in the IHOPE cohort were estimated, each of which included 20 households and approximately 100 individuals. Then, given the spatial location of each cluster, a raster surface of the whole district was obtained to improve visualization of results. This was done through inverse distance weighted interpolation on the empirical Bayes estimates of seroprevalence of each cluster. Second, the demographic, socio-economic and geographic factors associated with recent and past SARS-CoV2 infections were studied for Ifanadiana. Demographic factors were studied at the individual level and included age and sex. Socio-economic factors included individual’s occupation for those older than 15 years or school enrolment for those 5-14 years, as well as a household wealth index calculated through a principal components analysis of household assets, which was then split into five quintiles, following standard DHS methods [48]. Geographic factors were estimated at the cluster level and included the distance to major towns in the district (‘chef lieu de commune’ or commune capital), distance to the main paved road crossing the district from West to East, and distance to the secondary non-paved roads going north and south of the District from the main road. The Open Source Routing Machine (OSRM) engine was used to accurately estimate the shortest path between the villages in each cluster and these geographic features. For this, we had previously mapped the entire district of Ifanadiana on OpenStreetMap, resulting in over 20,000 km of footpaths and 5000 residential areas mapped (see [49] for details). The average distance for each cluster was then split into two groups at 5km, a common threshold that represents approximately a 1h travel time on foot.

Associations for each of these factors were modelled individually using univariate and multivariate logistic regressions in generalized linear models. Sampling weights that adjusted for unequal probability of selection due to stratification and non-response were calculated for household surveys. Estimates (totals, proportions, odds ratios) were obtained using survey commands available in R-package *survey* and applicable sampling weights [50]. All analyses were done for each individual serological marker and for the composite indicators based on the PCA and k-means clustering. All analyses were performed with R software, version 4.2.1 [51], and R packages “survey”, “gstat”, “rgdal”, “stats”, and “ggplot2”.

#### Excess mortality and infection fatality rates associated with SARS-CoV2

Adult mortality was estimated from the IHOPE cohort using the synthetic life-table method for DHS surveys [48]. First, six-year averages of adult mortality per 1000 population, split by age group and sex, were estimated for each wave of the cohort in order to obtain robust estimates over time that are comparable to standard DHS methods, before and after the COVID-19 epidemic. Because the period for these six-year estimates overlaps (surveys were repeated every two years from 2014 to 2021), mortality rates per year were then estimated from the 2021 wave of the cohort only, for the ten years prior to the survey. From this, a ten-year average was estimated (2012-2021) and excess mortality occurring in the years 2020 and 2021 was estimated as the difference between these years’ mortality and the ten-year average. Not all excess mortality in 2020-2021 can be assumed to be directly the result of COVID-19 deaths. To estimate expected excess mortality associated directly with SARS-CoV2 infections in our population, infection fatality rate (IFR) estimates per year of age (including lower and upper bounds for these estimates) were obtained from a recent study by the COVID-19 Forecasting Team [52], which are based on over 3000 surveys from dozens of countries. These age-specific IFR values were then combined with the observed age-specific number of SARS-CoV2 cases and age distribution in our population to obtain an expected excess mortality by age group. Observed excess mortality in our cohort was then compared with expected excess mortality for our population based on these IFR values.

## Results

### SARS-CoV-2 seroprevalence trends and associated factors

Overall, 6496 individuals were included in the seroprevalence analyses, nearly half of whom were children under 15 years, with an even sex ratio (Table 1). Only one individual out of ten individuals reported COVID-related symptoms in the 6 months prior to the survey, with the most common symptoms being fever (6.5%) and respiratory problems (4.1%). Despite low prevalence of reported symptoms, district seroprevalence rates were substantially higher, ranging from 5.1% (Spike RBD) to 43.8% (Spike S2) for IgG antibodies, and from 7.2% (Spike S1) to 17.9% (Spike S2) for IgM antibodies. Positivity to Spike S2 was higher than to any of the other markers, both for IgG and IgM antibodies. After clustering via PCA and k-means, overall seroprevalence was estimated to be 18%, with 10.1% having a predominantly IgG response suggestive of past infection and 7.9% having a predominantly IgM response suggestive of recent infection.

**Table 1.**
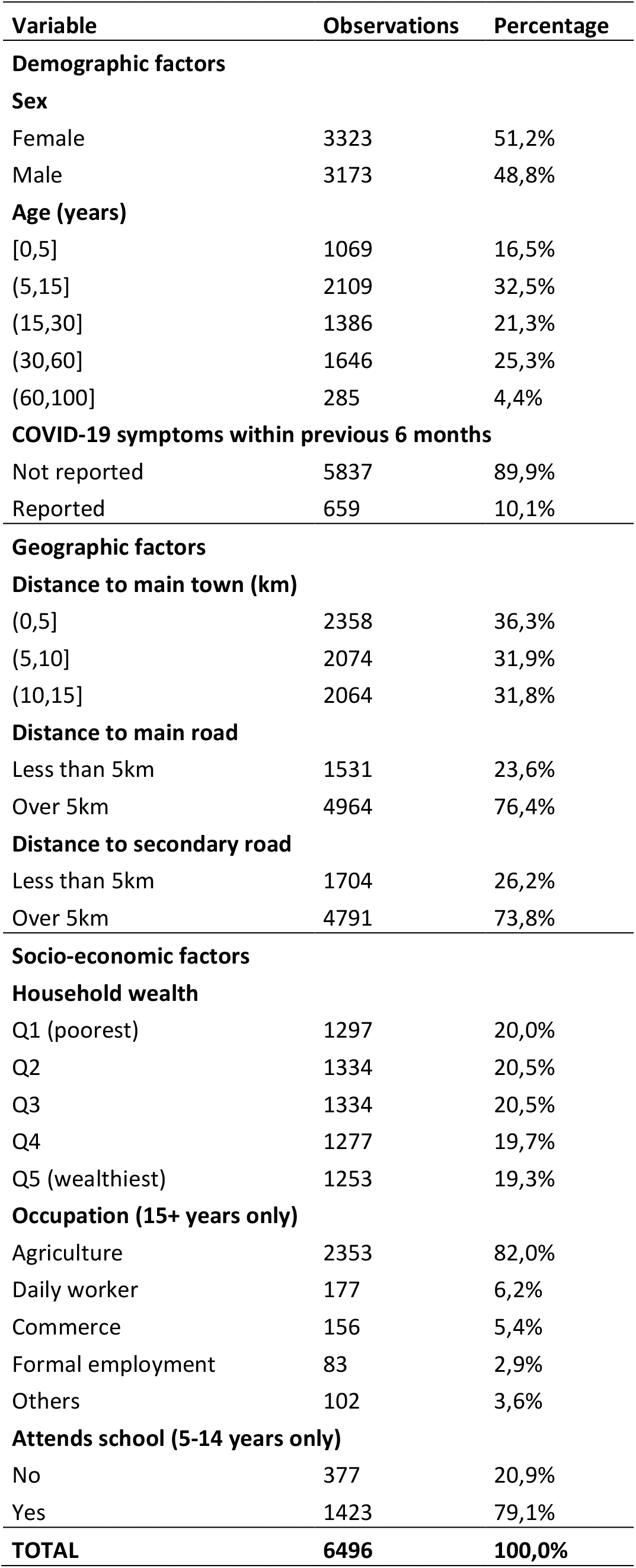
Characteristics of individuals included in the COVID-19 seroprevalence survey

| **Variable** | **Observations** | **Percentage** |
| --- | --- | --- |
| **Demographic factors** |  |  |
| **Sex** |  |  |
| Female | 3323 | 51,2% |
| Male | 3173 | 48,8% |
| **Age (years)** |  |  |
| [0,5] | 1069 | 16,5% |
| (5,15] | 2109 | 32,5% |
| (15,30] | 1386 | 21,3% |
| (30,60] | 1646 | 25,3% |
| (60,100] | 285 | 4,4% |
| **COVID-19 symptoms within previous 6 months** |  |  |
| Not reported | 5837 | 89,9% |
| Reported | 659 | 10,1% |
| **Geographic factors** |  |  |
| **Distance to main town (km)** |  |  |
| (0,5] | 2358 | 36,3% |
| (5,10] | 2074 | 31,9% |
| (10,15] | 2064 | 31,8% |
| **Distance to main road** |  |  |
| Less than 5km | 1531 | 23,6% |
| Over 5km | 4964 | 76,4% |
| **Distance to secondary road** |  |  |
| Less than 5km | 1704 | 26,2% |
| Over 5km | 4791 | 73,8% |
| **Socio-economic factors** |  |  |
| **Household wealth** |  |  |
| Q1 (poorest) | 1297 | 20,0% |
| Q2 | 1334 | 20,5% |
| Q3 | 1334 | 20,5% |
| Q4 | 1277 | 19,7% |
| Q5 (wealthiest) | 1253 | 19,3% |
| **Occupation (15+ years only)** |  |  |
| Agriculture | 2353 | 82,0% |
| Daily worker | 177 | 6,2% |
| Commerce | 156 | 5,4% |
| Formal employment | 83 | 2,9% |
| Others | 102 | 3,6% |
| **Attends school (5-14 years only)** |  |  |
| No | 377 | 20,9% |
| Yes | 1423 | 79,1% |
| **TOTAL** | **6496** | **100,0%** |

**Table 2.** Reported COVID-19 symptoms in the previous 6 months and estimated COVID-19 seroprevalence, all ages (N=6496)

| **COVID-19 symptom (last 6 months)** | **Percentage (95% CI)** |
| --- | --- |
| Fever | 6.46 (5.3-7.6) |
| Cough or resp. problems | 4.1 (3.2-5.0) |
| Headache | 3.9 (2.9-4.8) |
| Fatigue or pain in muscles/joints | 3.3 (2.5-4.0) |
| Diarrhea or nausea | 1.1 (0.8-1.5) |
| Loss of smell | 0.8 (0.4-1.2) |
| Sore throat | 0.4 (0.2-0.6) |
| **SARS Cov-2 serological marker** | **Seroprevalence (95% CI)** |
| **IgG** |  |
| Spike S1 | 13.0 (10.6-15.4) |
| Spike S2 | 43.8 (40.9-46.7) |
| Spike RBD | 5.1 (3.8-6.3) |
| Spike NP | 16.4 (14.1-18.7) |
| **IgM** |  |
| Spike S1 | 7.2 (6.3-8.2) |
| Spike S2 | 17.9 (15.8-19.9) |
| Spike RBD | 9.1 (7.9-10.3) |
| Spike NP | 9.1 (7.5-10.7) |
| **Composite (k-means clustering)** |  |
| Infected vs. healthy | 18.0 (15.9-20.1) |
| IgG Predominance (past infection) | 10.1 (8.3-11.9) |
| IgM Predominance (recent infection) | 7.9 (6.5-9.2) |

The spatial distribution of past infections in Ifanadiana suggests that COVID-19 cases during the first wave accumulated predominantly in close proximity to the paved road, with the exception of a few clusters in remote areas in the north of the district where seroprevalence reached nearly 30% (Figure 2). In contrast, recent infections during the second wave were more evenly distributed across the district, with very low prevalence in clusters located along the main road. Similarly, the factors associated with SARS-CoV-2 seroprevalence varied substantially between past and recent infections (Figure 3 and Table 4). In both recent and past infections, seroprevalence was similar for males and females, increased with age and decreased with distance to a major town. However, seroprevalence for past infections was higher for individuals in the wealthiest household quantile and those whose occupation was not agriculture (e.g. commerce, formal employment, etc.), with opposite associations for recent infections. In addition, seroprevalence of recent infections doubled for those individuals reporting a COVID-19 symptom in the previous 6 months compared to those not reporting symptoms, while no association was observed for past infections. Spatial distributions and associations for each of the eight SARS-CoV-2 serological markers are available in the Appendix, Section S1.

**Table 4.** Logistic regression results for associations with SARS-CoV-2 seroprevalence in past and recent infections, based on IgG and IgM predominance

| Variable | IgG (Past infection) |  | IgM (Recent infection) |  |
| --- | --- | --- | --- | --- |
|  | Univariate Odds Ratio (95% CI) | Adjusted Odds Ratio (95% CI) | Univariate Odds Ratio (95% CI) | Adjusted Odds Ratio (95% CI) |
| **Demographic factors** |  |  |  |  |
| **Sex (ref. female)** |  |  |  |  |
| Male | 0.95 (0.79-1.14) | - | 0.82 (0.64-1.05) | - |
| **Age in years (ref. 0-5)** |  |  |  |  |
| (5,15] | 0.97 (0.74-1.28) | - | 2.17 (1.32-3.58)** | 2.24 (1.36-3.71)** |
| (15,30] | 0.94 (0.71-1.24) | - | 3.32 (1.97-5.59)*** | 3.45 (2.04-5.85)*** |
| (30,60] | 1.21 (0.9-1.62) | - | 4.12 (2.51-6.75)*** | 4.07 (2.48-6.68)*** |
| (60,100] | 1.25 (0.86-1.81) | - | 4.64 (2.38-9.04)*** | 4.34 (2.22-8.48)*** |
| **COVID-19 symptoms (ref. not reported)** |  |  |  |  |
| Reported | 1.03 (0.75-1.42) | - | 2.05 (1.35-3.12)** | 1.91 (1.26-2.91)** |
| **Geographic factors** |  |  |  |  |
| **Distance to health center (ref. 0-5km)** |  |  |  |  |
| (5,10] | 0.72 (0.44-1.17) | - | 1.12 (0.72-1.75) | 1.21 (0.86-1.7) |
| (10,15] | 0.74 (0.45-1.23) | - | 0.75 (0.47-1.18) | 0.74 (0.52-1.05). |
| **Distance to main road (ref. 0-5km)** |  |  |  |  |
| Over 5km | 0.55 (0.37-0.81)** | 0.7 (0.49-1.02). | 1.18 (0.64-2.19) | - |
| **Distance to secondary road (ref. 0-5km)** |  |  |  |  |
| Over 5km | 0.69 (0.43-1.11) | - | 0.67 (0.45-1). | 0.62 (0.44-0.86)** |
| **Socio-economic factors** |  |  |  |  |
| **Household wealth (ref. Q1 - poorest)** |  |  |  |  |
| Q2 | 1.07 (0.74-1.57) | 1.04 (0.71-1.53) | 0.84 (0.56-1.27) | 0.79 (0.53-1.17) |
| Q3 | 1.03 (0.69-1.54) | 0.97 (0.65-1.45) | 0.93 (0.64-1.35) | 0.84 (0.6-1.17) |
| Q4 | 1.37 (0.89-2.12) | 1.23 (0.79-1.92) | 1 (0.65-1.54) | 0.83 (0.56-1.22) |
| Q5 (wealthiest) | 2.14 (1.4-3.27)*** | 1.69 (1.14-2.52)* | 0.67 (0.31-1.44) | 0.48 (0.26-0.88)* |
| **Occupation, individuals 15+ years (ref. Agriculture)<sup>1</sup>** |  |  |  |  |
| Daily worker | 1.31 (0.78-2.18) | - | 0.71 (0.39-1.31) | - |
| Commerce | 2.03 (1.13-3.67)* | - | 1.14 (0.22-5.97) | - |
| Formal employment | 1.78 (1.04-3.07)* | - | 0.77 (0.36-1.65) | - |
| Others | 1.74 (0.87-3.47) | - | 0.35 (0.12-1.01). | - |
| **Attends school, children 5-14 years (ref. No)<sup>1</sup>** |  |  |  |  |
| Yes | 1.29 (0.8-2.06) | - | 0.65 (0.44-0.97)* | - |

. $p < 0.1$ ; \* $p < 0.05$ ; \*\* $p < 0.01$ ; \*\*\* $p < 0.001$
<sup>1</sup> Variable only applicable to a population subgroup, not included in multivariate analyses

**Figure 2.**
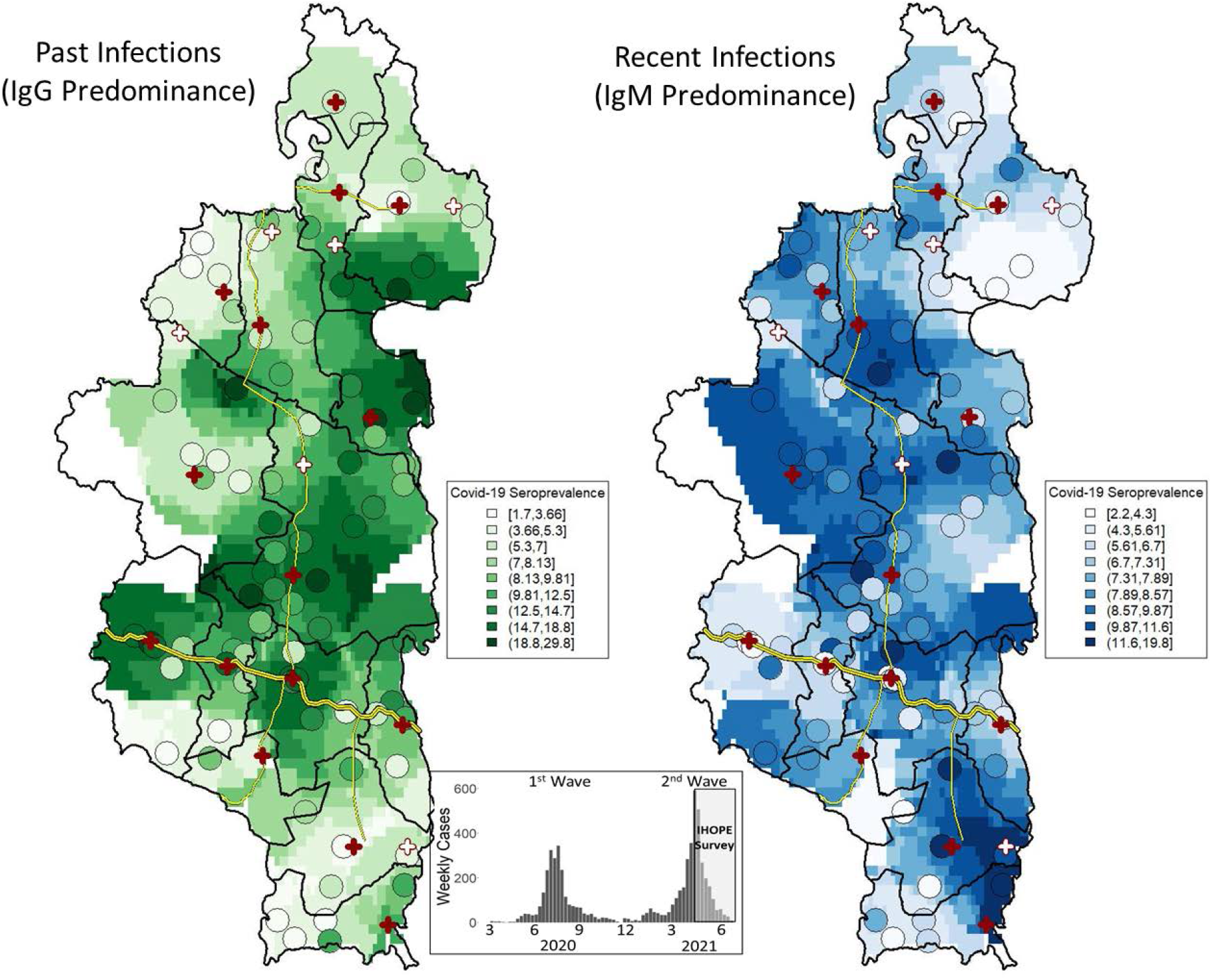
Spatial distribution of SARS-CoV-2 seroprevalence in Ifanadiana District. Left map shows seroprevalence of past infections (IgG predominance in k-clustering) and right map shows seroprevalence of recent infections (IgM predominance), with colours ranging from light (low seroprevalence) to dark (high seroprevalence). Average seroprevalence and location of each of the 80 clusters in the survey are represented by circles, while the rest of the raster is based on inverse distance weighted interpolation. Bottom inset figure shows the timing of the survey with respect to the COVID-19 epidemic in Madagascar.

**Figure 3.**
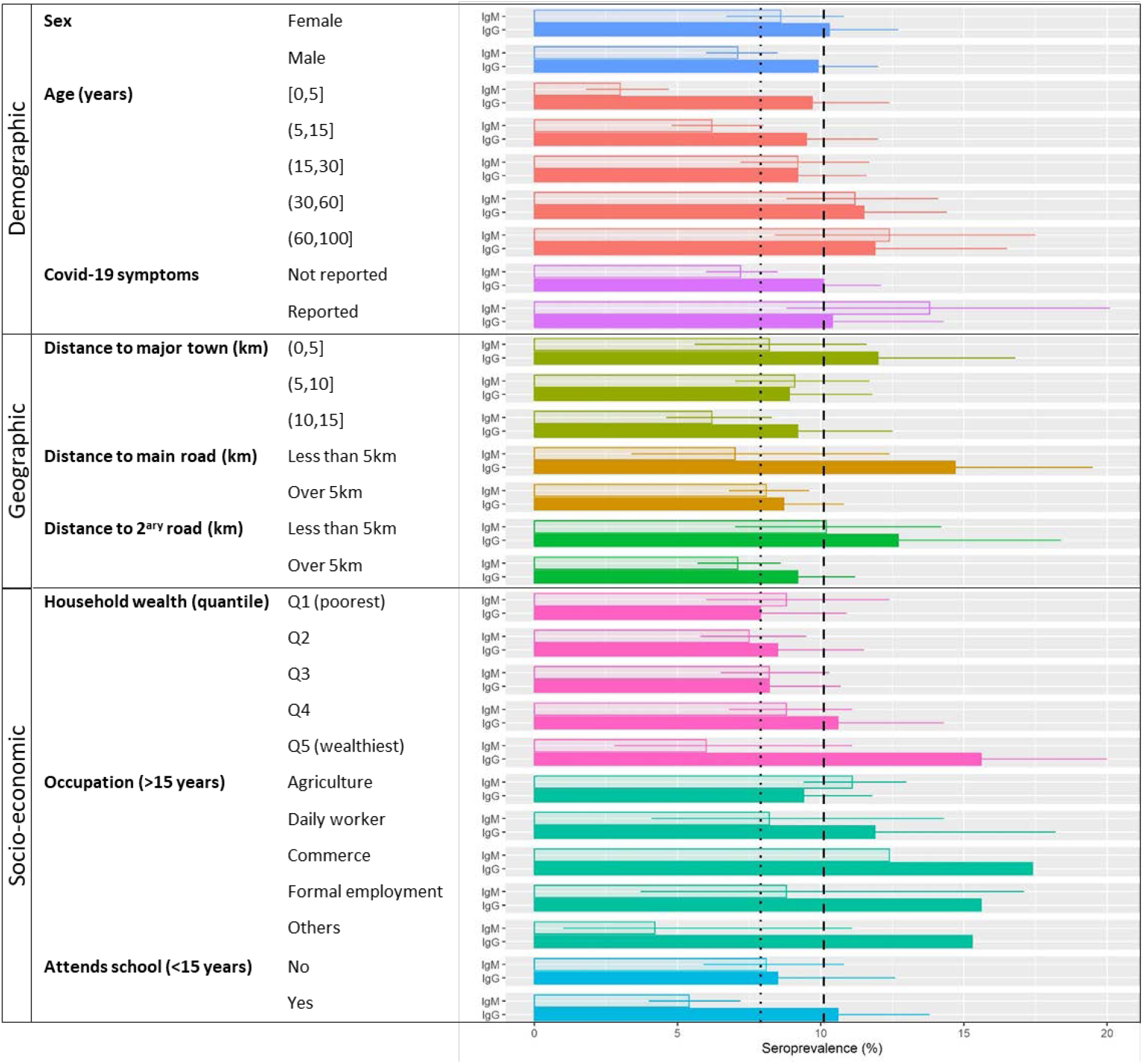
Factors associated with SARS-CoV-2 seroprevalence in Ifanadiana District. Horizontal bars show average seroprevalence per group, split into past infections (IgG, filled colour bars) and recent infections (IgM, translucent colour bars), with 95% confidence intervals as whiskers. Vertical lines represent average seroprevalence in Ifanadiana for past infections (dashed) and recent infections (dotted). Four confidence interval limits (Occupation variable) were removed to improve visualisation of results.

### Mortality and infection fatality rates associated with COVID-19

Estimates of 6-year adult mortality in the 2021 wave of the IHOPE cohort collection were higher than in all previous waves of data collection (Figure 4). The largest increase was observed for older men (35-49 years), who experienced a nearly 100% increase in mortality in reports from the 2021 cohort wave as compared to the 2018 cohort wave (from 10.6 to 19.3 per 1000). For women and young men, mortality had been slowly but steadily declining in the previous three waves, and then increased by 20-50% in the 2021 cohort wave to a level higher than at the 2014 baseline. Similar trends were observed using data from the 2021 cohort wave only, where analyses of annual mortality rates in the previous 10 years showed that mortality in 2020 was substantially higher than average, especially for older men (Figure 4). However, these time series were more stochastic given lower sample sizes and, for women and young men, the peak in mortality observed in 2020 was not higher than other peaks observed in previous years. Overall, excess mortality for 2020-2021 was estimated at 1.61 per 1000 for individuals 15-34 years and 4.82 per 1000 for individuals 35-49 years. Given known COVID-19 IFRs and observed seroprevalence by age group, expected excess mortality associated directly with COVID-19 infections in our cohort would be 0.05 (range 0.04-0.08) per 1000 for individuals 15-34 years and 0.54 (range 0.39-0.85) per 1000 for individuals 35-49 years (Table 5). This suggests that if IFRs in Ifanadiana are consistent with those previously estimated, only 3.1% (range 2.5-5.0%) and 8.9% (range 6.5-14.1%) of observed excess mortality for individuals 15-34 years and 35-49 years, respectively, would be directly associated with COVID-19 deaths.

**Table 5.** Estimation of excess adult mortality and percentage that could be directly attributed to COVID-19 given observed seroprevalence and known infection fatality ratios by age.

| Age Group | Seroprevalence (%) | Observed excess mortality 2020-2021 (per 1000) | Expected excess mortality directly from COVID-19 infections (per 1000) | % of excess mortality directly attributed to COVID-19 infections |
| --- | --- | --- | --- | --- |
| 15-34 Years | 32.7 | 1.61 | 0.05 (0.04-0.08) | 3.1 (2.5-5.0) |
| 35-49 Years | 35.8 | 6.04 | 0.54 (0.39-0.85) | 8.9 (6.5-14.1) |

**Figure 4.**
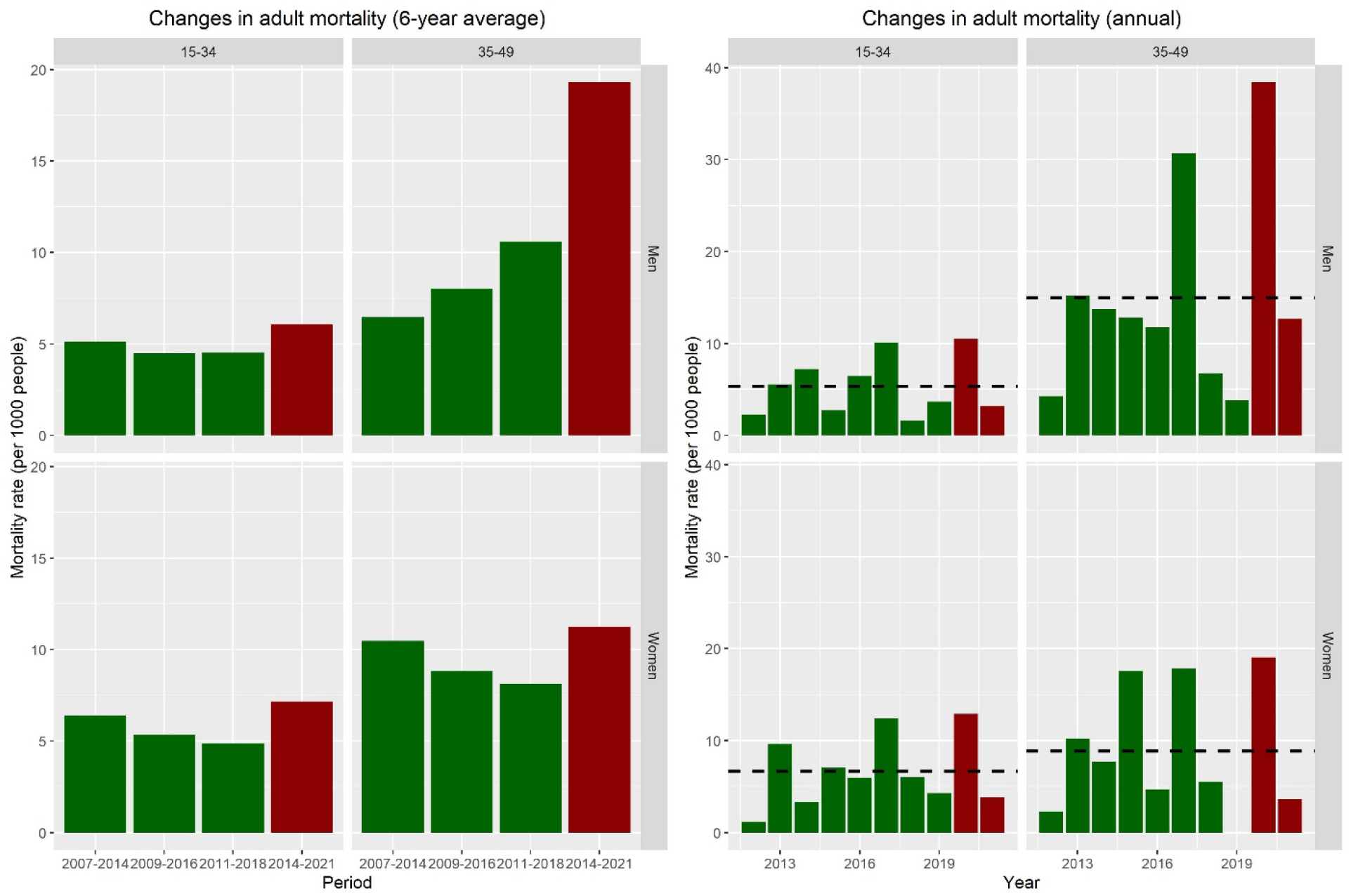
Trends in adult mortality rates in the IHOPE cohort, Ifanadiana District. Graphs show mortality per 1000 people before (green) and during (red) the COVID-19 pandemic. Left panels show changes in mortality rates across survey waves using the 6-year average prior to each survey wave. Right panels show changes in mortality rates per year in the 10 years prior to the 2021 survey. Dashed lines are the 10-year average for the period for each age and sex group. Note that the 2021 survey was conducted in April-June, so mortality estimates for this year only comprise part of the year.

## Discussion

Nearly three years after the start of the COVID-19 pandemic, the burden of the disease in areas that are traditionally most vulnerable - rural populations in the developing world - is still unclear. SARS-CoV-2 seroprevalence surveys have been essential for a better understanding of COVID-19 transmission across the world and among particular populations, but high quality studies have rarely focused on rural areas of sub-Saharan Africa [13,52]. Using a population-representative longitudinal cohort and a nested seroprevalence survey of nearly 6500 people of all ages, we provide a fine-scale account of COVID-19 spread and burden in a rural district of Madagascar during its first two epidemic waves. Our results suggest that despite low density and connectivity in the majority of the district, about one in five people had been infected by the period of April-June 2021. COVID-19 primarily spread along the only paved road in the district during the first wave, and then spread along secondary roads during the second wave to more remote areas in the north and south of the district. Adult mortality increased in 2020, particularly for older men, but the majority of the excess mortality during this period could not be directly attributed to COVID-19 deaths given known infection fatality ratios and observed seroprevalence in the area. This suggests that for populations living in rural, low-resource settings, COVID-19 could have significant health impacts, either because of higher IFRs than previously estimated or because of substantial indirect impacts on health care.

Our results reveal that seroprevalence was lower in this rural district of Madagascar than in the nearby city of Fianarantsoa (about 1h30 drive from Ifanadiana), where seroprevalence in blood donors was already about 20% by November 2020 [15] and over 40% by February-June 2021 according to a cross-sectional household survey [53]. This is consistent with studies conducted in other African settings, which found that seroprevalence tends to be lower in rural populations than in their urban counterparts [13,54–60], with a few exceptions [61,62]. However, our study goes further, revealing the substantial heterogeneity in the spatio-temporal patterns of rural infections. Populations in the district living within 5km to roads or large towns had comparable or higher seroprevalence rates to those from urban Fianarantsoa [15,53], which highlights the major role played by population density and road connectivity in the spread of COVID-19 even in rural areas of the developing world where both factors are significantly lower than in urban settings.

Seroprevalence rates differed across demographic and socio-economic groups in our population. Seroprevalence in both epidemic waves increased with age, especially for those older than 30, who had higher risk of infection than young children, but risk did not differ between men and women. Although associations with demographic factors can be context-specific and vary across settings, a recent review of seroprevalence studies found similar average trends for Africa [13]. Socio-economic factors also modified individuals’ risk of infection in this largely impoverished population, where the primary occupation is subsistence agriculture. Wealthier individuals and those with certain occupations such as commerce and formal employment were at higher risk of being infected in the first wave but at lower risk of being infected in the second. It is well known that individuals with high social connectivity are at higher risk of infection and can contribute disproportionately to the spread of diseases such as COVID-19 [63–65]. While research on at-risk occupational activities in Africa has mostly focused on healthcare workers [25], a better understanding of the role played by other socio-economic groups with high mobility and social connections on COVID-19 spread could open new possibilities for disease control [66].

In this poor rural setting where mortality rates were already high prior to the pandemic, our results suggest that the COVID-19 epidemic was associated with a substantial increase in adult mortality. The increase observed for Ifanadiana was similar to that found in Sudan’s capital [67], where a 67% rise was observed in a cross-sectional survey. However, about three quarters of deaths in the Sudan study were among individuals 50 years or older, a vulnerable population group that was not assessed here due to study design limitations. The excess mortality in individuals aged 15-49 years in Ifanadiana was substantially higher than what could have been expected based on observed seroprevalence and known infection fatality rates for these age groups in other parts of the world [52], which would only explain a small fraction of excess mortality. Possible explanations are that i) other factors unrelated to COVID-19 were associated with higher mortality in this period; ii) IFRs in our population were higher than those estimated from other settings; and/or iii) COVID-19 had indirect impacts on mortality, such as through the effects on healthcare access and health system disruptions. Interestingly, the only other outlying year for mortality in this ten-year period was 2017, when Madagascar’s largest plague epidemic in recent history occurred [68].

Our study had several limitations. First, although we used robust clustering methods with information on both IgG and IgM for multiple antibodies against SARS-CoV-2 to classify infections into past and recent, there is uncertainty around such classification. For instance, IgM titres can remain high in past symptomatic infections and IgG NP titres can increase early in the infection [44,69]. However, the fact that the first epidemic wave occurred nearly a year prior and that results of associated factors in each wave are consistent with known patterns of COVID-19 spread (e.g. initial spread along better connected populations) suggests that potential misclassification biases had little impact in the overall analysis. Second, low sample sizes for analyses of adult mortality in our cohort could have impacted our estimates of excess mortality, especially because annual rates in the 10-year period were not stable. Despite this, results of these analyses were consistent with those based on 6-year averages for each wave of the cohort, both in the relative increase in mortality and the demographic groups most affected. Third, our survey mirrored a DHS design, where mortality estimates are based on information for siblings provided in interviews with men and women of reproductive age. As a result, even though individuals older than 50 years are the most likely to be affected, we could not assess the impact of COVID-19 on this age group due to low sample sizes. Fourth, the survey was conducted in the middle of the second wave of COVID-19, which prevented us from fully capturing the impact of this wave. This could also have affected the trends observed for Ifanadiana if the timing of the survey in different parts of the district had an impact on their corresponding seroprevalence. However, a complementary analysis of seroprevalence over survey dates suggests this was unlikely to be the case (Figure S9). Finally, as is the case with any local-scale survey, the results of this study do not necessarily represent the COVID-19 situation in other parts of Madagascar or sub-Saharan Africa.

In conclusion, our study provides an unusually detailed picture of COVID-19 morbidity and mortality in a poor rural setting of sub-Saharan Africa and suggests that the disease burden in these areas may have been substantially underestimated. The overall lack of diagnostic capacity, representative seroprevalence studies and routine systems to appropriately capture and analyse COVID-19 infections and deaths in these settings may have contributed to a misperception that sub-Saharan Africa has been largely spared from this pandemic. Given known vulnerabilities of these populations to other infectious diseases, combined with the fragility of their health systems, more attention and quality research is needed to better understand the true burden of COVID-19 in poor rural areas of sub-Saharan Africa and to devise appropriate responses to this and future pandemics.

## Data Availability

All data produced in the present study are available upon reasonable request to the authors

## ACKNOWLEDGEMENTS

The authors acknowledge the contributions and support of Felana Ihantamalala and Mauricianot Randriamihaja. They are grateful to all of the staff at PIVOT for their field support and their remarkable work in strengthening the health system in Ifanadiana. Thanks are due to the Madagascar Ministry of Health, at both the district and the central levels, for their continuous support and valuable insights. The authors also thank the Institut National de la Statistique (INSTAT) field teams for their involvement in the district-wide population survey.

## AUTHOR CONTRIBUTIONS

Conceived and designed the experiments: AG, ACM, KEF, MHB, MS. Performed laboratory analyses: LTR, MS. Performed statistical analyses: AG, ME. Contributed reagents/materials/data/analysis tools: AG, RJLR, ACM, MR, SA, LTR, MS. Wrote the initial draft of the manuscript: AG, MHB. Revised the manuscript and accepted it in its final form: AG, LTR, RJLR, ME, ACM, KEF, LFC, GC, BR, MR, SA, SP, RH, MHB, MS

## SUPPLEMENTARY INFORMATION

### Supplementary information Appendix

It contains 9 supplementary figures.

