## Supplemental information appendix for "Morbidity and mortality burden of COVID-19 in rural Madagascar: results from a longitudinal cohort and nested seroprevalence study"

- Supplementary Information Appendix -

Andres Garchitorena^1,2,3*^, Lova Tsikiniaina Rasoloharimanana^2^, Rado JL Rakotonanahary^3,4^, Michelle V Evans^1^, Ann C. Miller^4^, Karen E. Finnegan^3,4^, Laura F. Cordier^3^, Giovanna Cowley^3^, Benedicte Razafinjato^3^, Marius Randriamanambintsoa^5^, Samuel Andrianambinina^5^, Stephen Popper^6^, Raphaël Hotahiene^7^, Matthew H. Bonds^3,4✝^, Matthieu Schoenhals^2✝^

*^1^ MIVEGEC, Université de Montpellier, CNRS, IRD, Montpellier, France*

*^2^ Institut Pasteur de Madagascar, Antananarivo, Madagascar*

*^3^ NGO Pivot, Ifanadiana, Madagascar*

*^4^ Department of Global Health and Social Medicine, Harvard Medical School, Boston, MA, USA*

*^5^Direction de la Démographie et des Statistiques Sociales, Institut National de la Statistique, Antananarivo, Madagascar*

*^6^ Division of Infectious Diseases and Vaccinology, School of Public Health, University of California, Berkeley, CA, USA*

*^7^ Ministère de la Santé Publique, Antananarivo, Madagascar*

✝ Equal contribution

**Section S1. SARS-CoV-2 seroprevalence trends and factors associated for each serological marker**

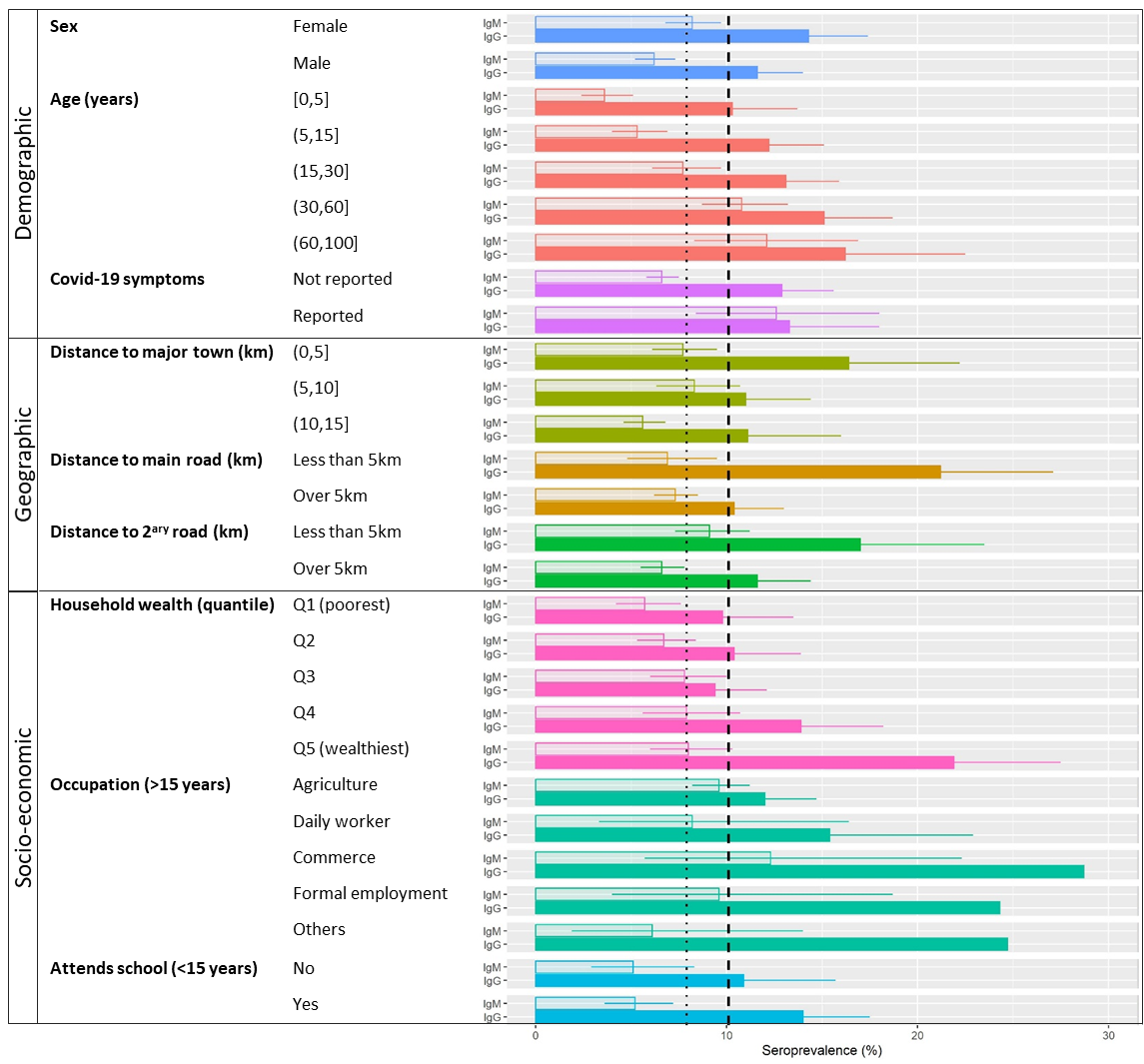

**Figure S1. Factors associated with SARS-CoV-2 Spike protein S1 seroprevalence in Ifanadiana District.** Horizontal bars show average seroprevalence per group, split into IgG (filled colour bars) and IgM (translucent colour bars), with 95% confidence intervals as whiskers. Vertical lines represent average seroprevalence in Ifanadiana for past infections (dashed) and recent infections (dotted). Some confidence interval limits were removed to improve visualisation of results.

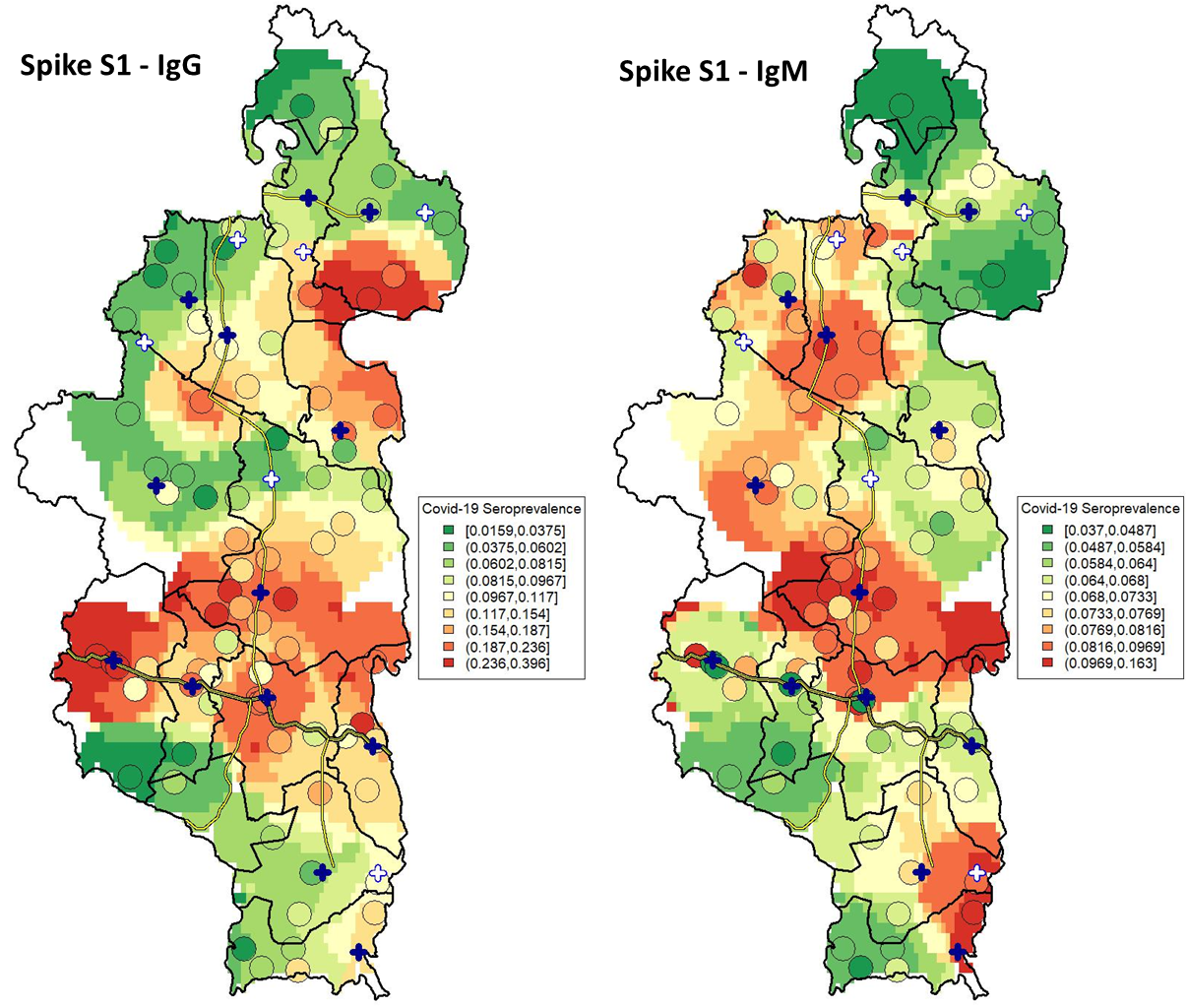

**Figure S2. Spatial distribution of seroprevalence to SARS-CoV-2 Spike protein S1 in Ifanadiana District.** Left map shows seroprevalence for IgG and right map shows seroprevalence for IgM, with colours ranging from green (low seroprevalence) to red (high seroprevalence). Average seroprevalence and location of each of the 80 clusters in the survey are represented by circles, while the rest of the raster is based on inverse distance weighted interpolation.

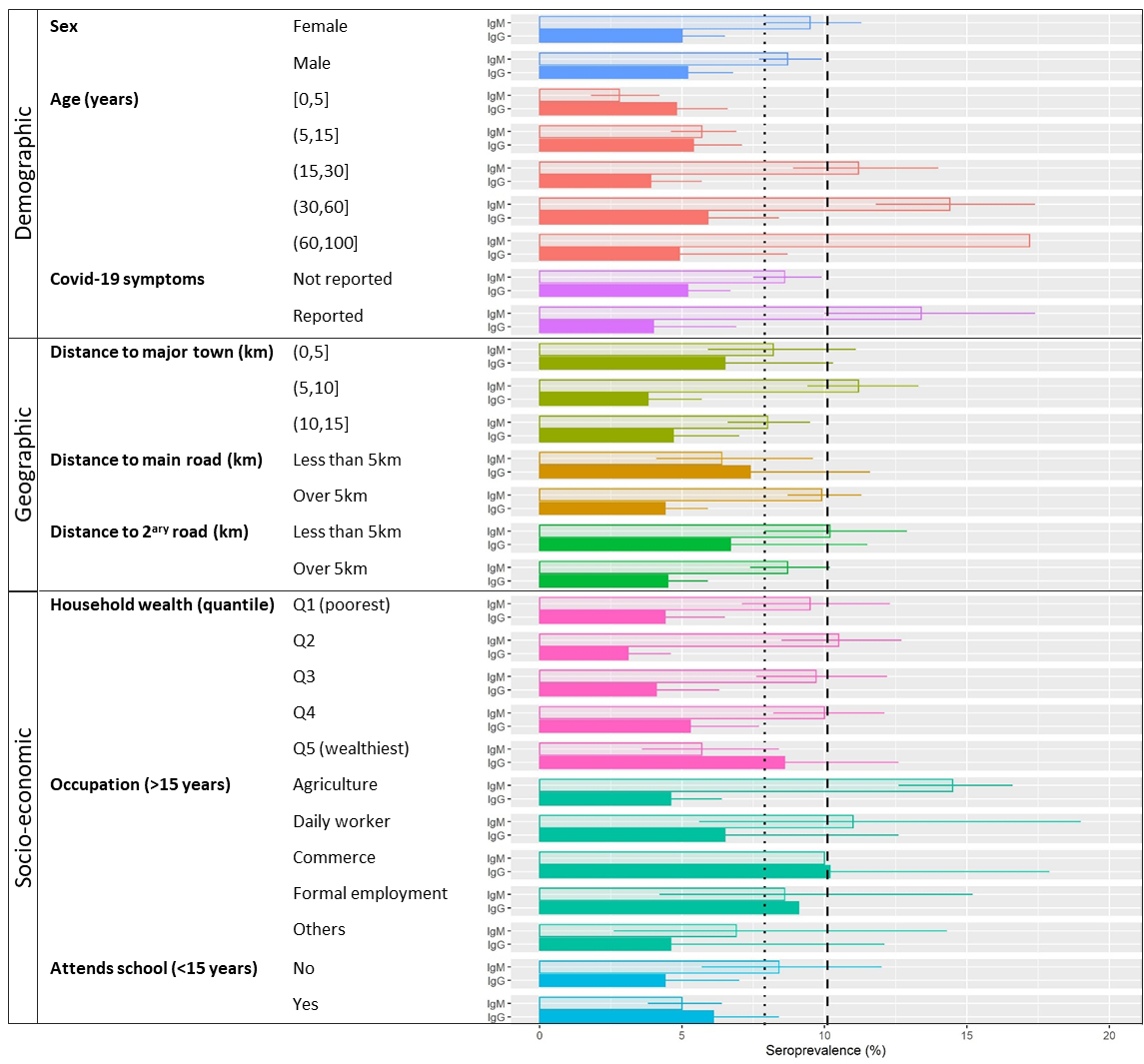

**Figure S3. Factors associated with SARS-CoV-2 Spike protein receptor-binding domain (RBD) seroprevalence in Ifanadiana District.** Horizontal bars show average seroprevalence per group, split into IgG (filled colour bars) and IgM (translucent colour bars), with 95% confidence intervals as whiskers. Vertical lines represent average seroprevalence in Ifanadiana for past infections (dashed) and recent infections (dotted). Some confidence interval limits were removed to improve visualisation of results.

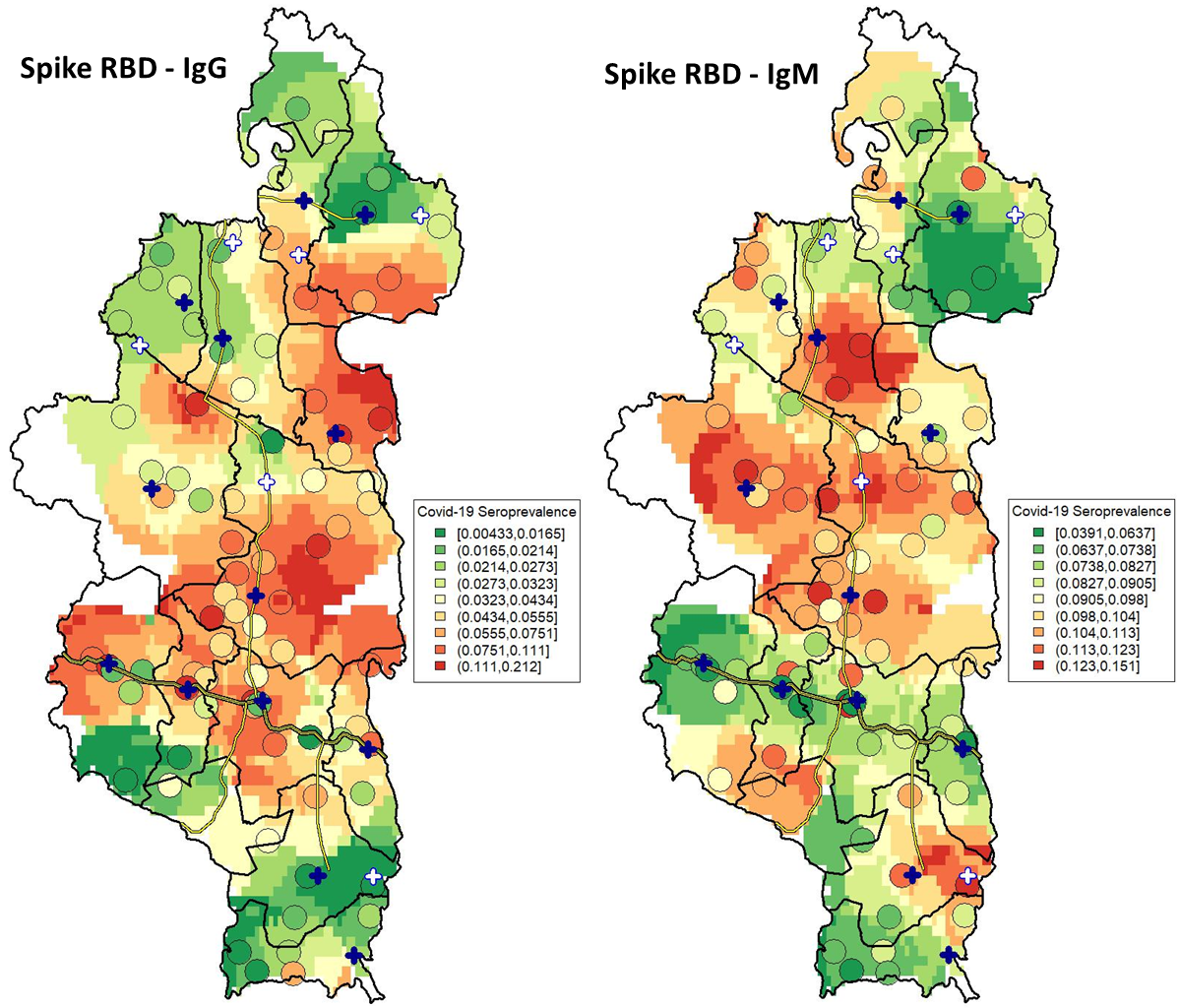

**Figure S4. Spatial distribution of seroprevalence to SARS-CoV-2 Spike protein receptor-binding domain (RBD) in Ifanadiana District.** Left map shows seroprevalence for IgG and right map shows seroprevalence for IgM, with colours ranging from green (low seroprevalence) to red (high seroprevalence). Average seroprevalence and location of each of the 80 clusters in the survey are represented by circles, while the rest of the raster is based on inverse distance weighted interpolation.

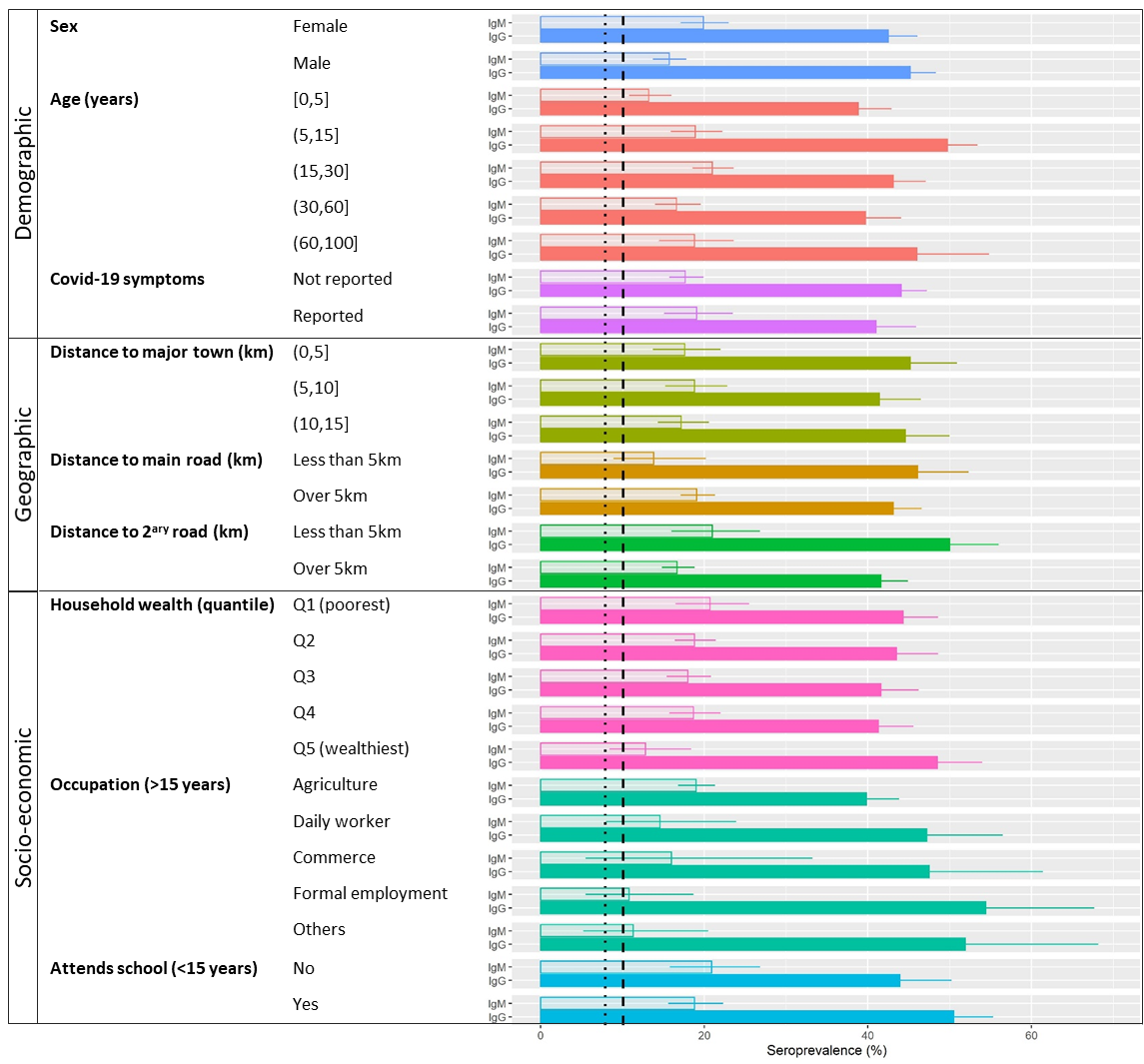

**Figure S5. Factors associated with SARS-CoV-2 Spike protein S2 seroprevalence in Ifanadiana District.** Horizontal bars show average seroprevalence per group, split into IgG (filled colour bars) and IgM (translucent colour bars), with 95% confidence intervals as whiskers. Vertical lines represent average seroprevalence in Ifanadiana for past infections (dashed) and recent infections (dotted). Some confidence interval limits were removed to improve visualisation of results.

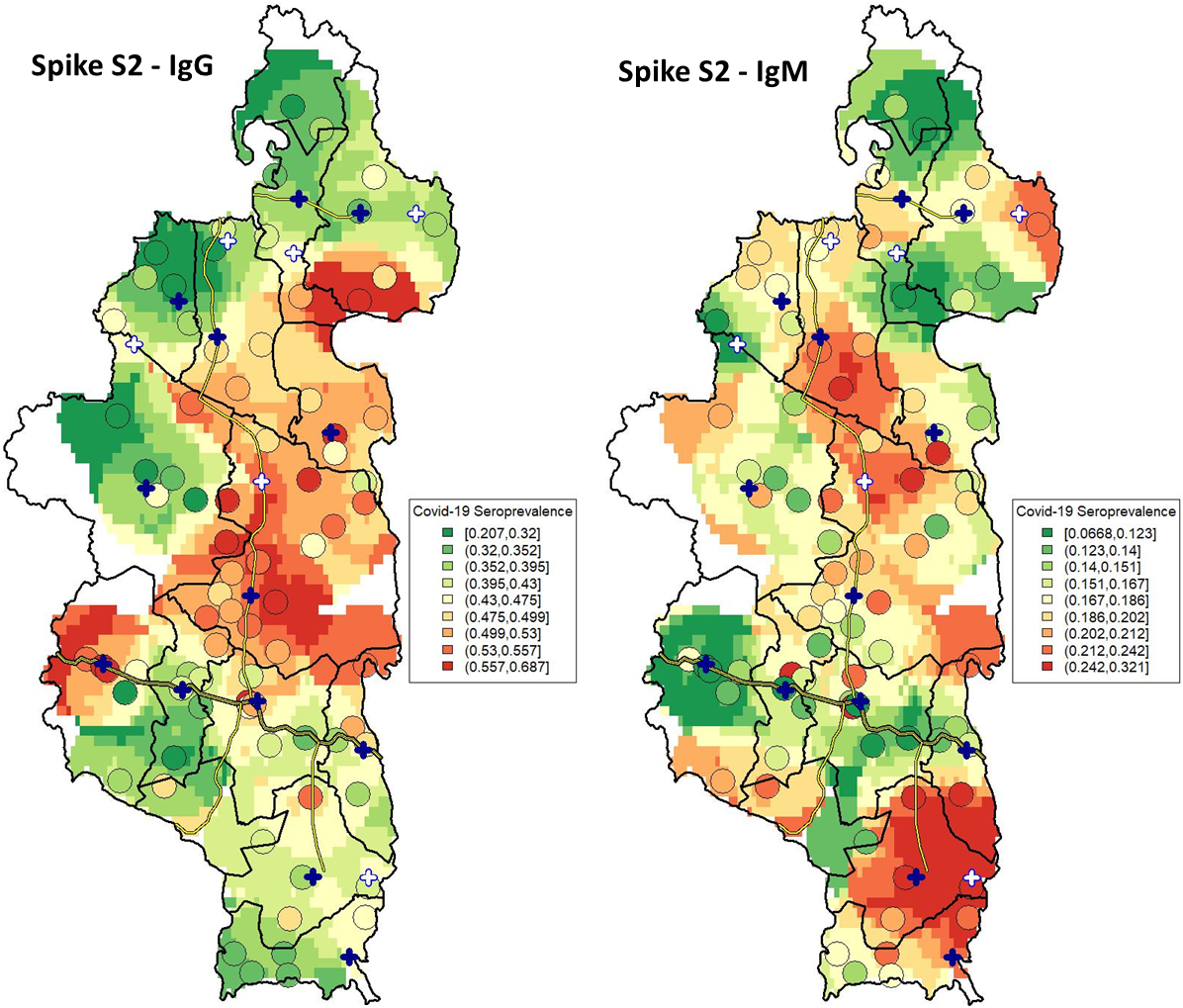

**Figure S6. Spatial distribution of seroprevalence to SARS-CoV-2 Spike protein S2 in Ifanadiana District.** Left map shows seroprevalence for IgG and right map shows seroprevalence for IgM, with colours ranging from green (low seroprevalence) to red (high seroprevalence). Average seroprevalence and location of each of the 80 clusters in the survey are represented by circles, while the rest of the raster is based on inverse distance weighted interpolation.

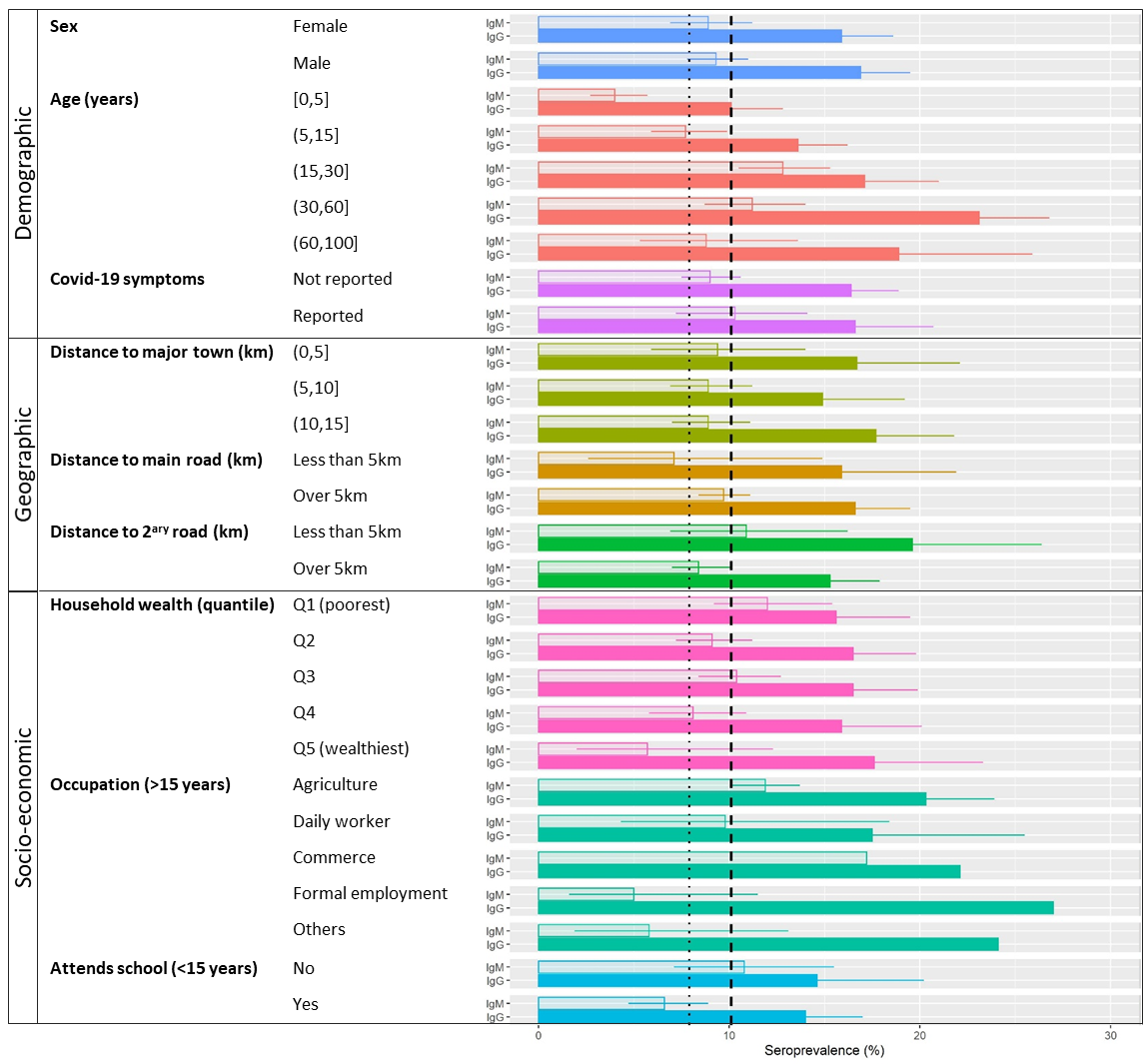

**Figure S7. Factors associated with SARS-CoV-2 Nucleocapsid protein (NP) seroprevalence in Ifanadiana District.** Horizontal bars show average seroprevalence per group, split into IgG (filled colour bars) and IgM (translucent colour bars), with 95% confidence intervals as whiskers. Vertical lines represent average seroprevalence in Ifanadiana for past infections (dashed) and recent infections (dotted). Some confidence interval limits were removed to improve visualisation of results.

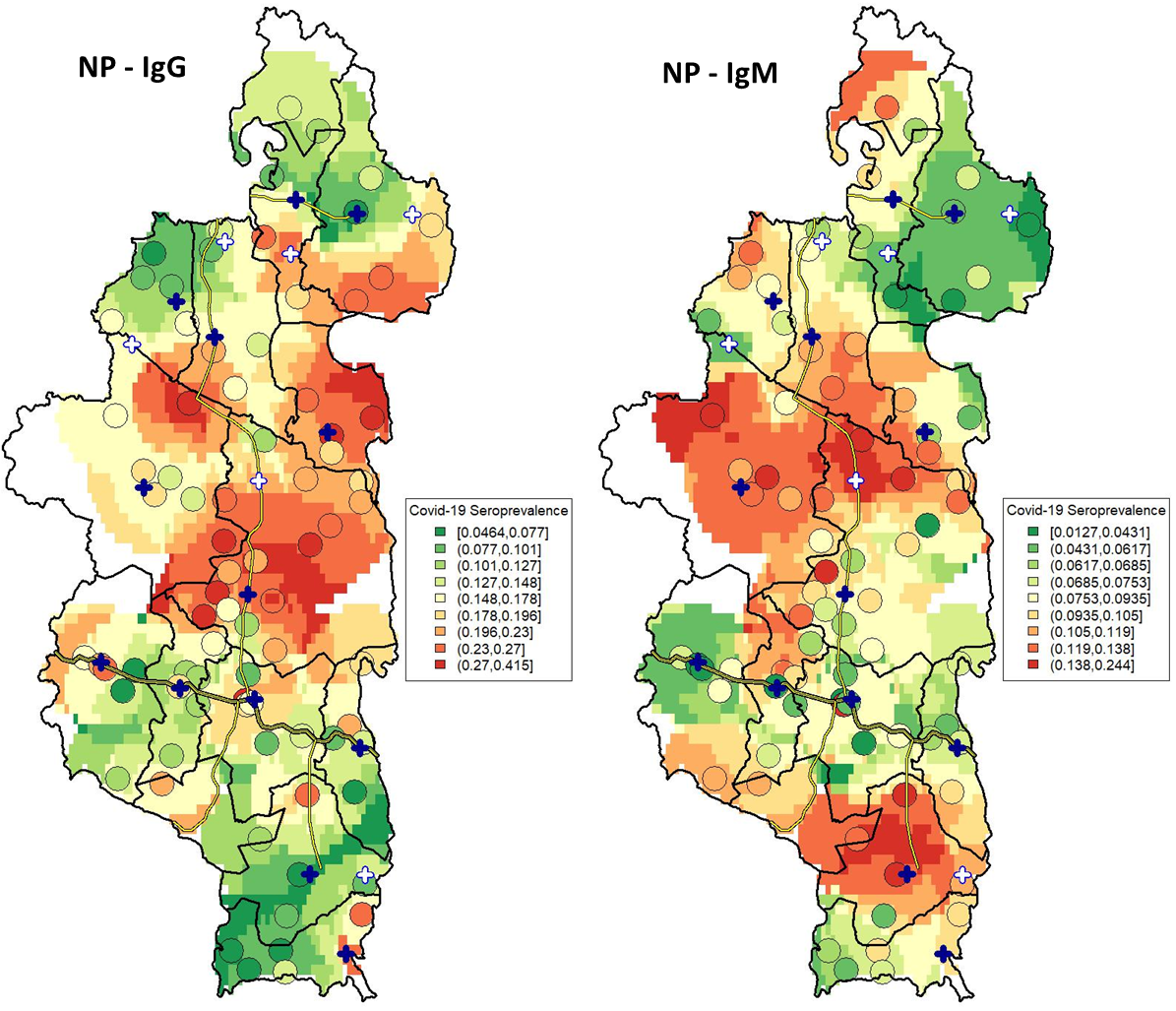

**Figure S8. Spatial distribution of seroprevalence to SARS-CoV-2 Nucleocapsid protein (NP) in Ifanadiana District.** Left map shows seroprevalence for IgG and right map shows seroprevalence for IgM, with colours ranging from green (low seroprevalence) to red (high seroprevalence). Average seroprevalence and location of each of the 80 clusters in the survey are represented by circles, while the rest of the raster is based on inverse distance weighted interpolation.

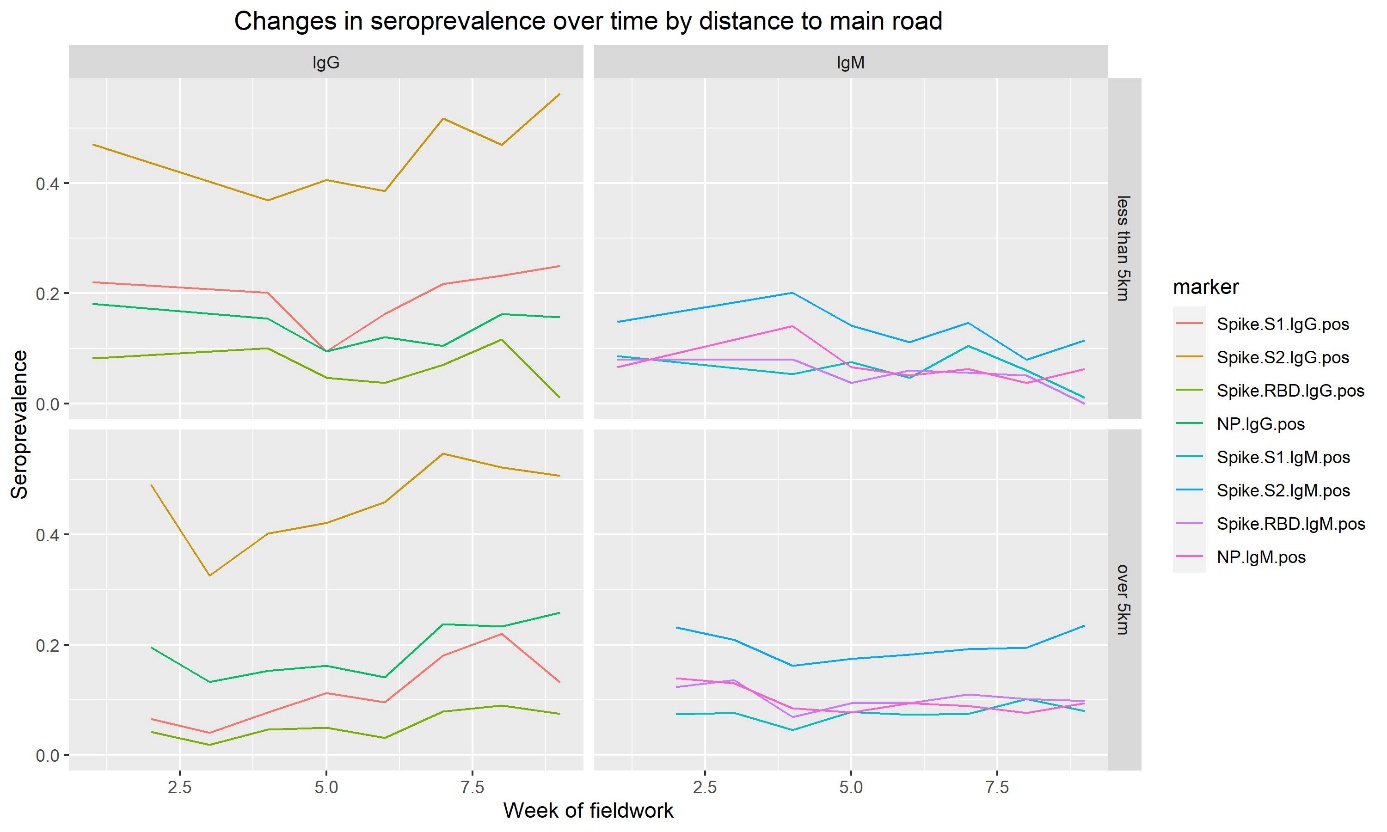

**Figure S9. Distribution of seroprevalence to SARS-CoV-2 in Ifanadiana District according to date of biological sample collection.** Left panel shows seroprevalence for IgG and right map shows seroprevalence for IgM, with top and bottom panels split according to cluster distance to the main road. Trends show the average seroprevalence for a particular week of fieldwork from April 22^nd^ to June 20^th^.
